# Predicting Unintended Pregnancy in Senegal by using Machine Learning Models: Evidence from Senegal DHS 2023

**DOI:** 10.1101/2024.11.24.24317850

**Authors:** Mushfiqur Rahman Khan Majlish, Sultanul Arafean Tawhid, Gulam Kibria, Nazmus Sakib

## Abstract

Unintended pregnancy refers to a pregnancy that is either mistimed or unwanted at the time of conception. Such pregnancies can have harmful effects, including negative impacts on maternal and child health, economic hardship, and strained relationships. This study aims to assess the effectiveness of machine learning algorithms in forecasting unintended pregnancies in Senegal and identifying the key factors that significantly impact them. The study utilized data from the 2023 Senegal Demographic and Health Survey, focusing on pregnant women. A final sample of 885 respondents was analyzed after handling missing values and six machine learning models namely Logistic Regression, Random Forest, K-Nearest Neighbors, Support Vector Machine, Naïve Bayes and Extra Tree Classifier was used. The Random Forest algorithm emerged as the best predictive model due to its highest AUC value (80.45%), surpassing all other machine learning algorithms used in this study. Total birth, currently residing with husband/partner, respondent’s education level, number of living children, Husband/partner’s occupation, residence type, Respondent can refuse sex and intention of contraceptive use are identified as contributing factors in that predict unintended pregnancy. The study’s results suggest that machine learning models, particularly Random Forest, can significantly enhance predictive accuracy for unintended pregnancies, helping public health initiatives in Senegal target at-risk populations. By identifying women at risk of unplanned pregnancies, targeted interventions and support services can be implemented, ultimately improving maternal health outcomes.

## Introduction

An unintended pregnancy is defined as a pregnancy that occurs when it is either unwanted (when no more children are desired) or mistimed (occurring earlier than desired). This classification highlights the distinction between pregnancies that are not planned at all and those that happen sooner than expected [1]. Globally, between 2010 and 2014, approximately 44% of all pregnancies were considered unplanned. In developed regions, the incidence of unintended pregnancies saw a significant decline of 30%, dropping from 64 per 1,000 women aged from 15 to 44 in 1990 to 1994 to 45 in 2010 to 2014. Conversely, in developing regions, the decline was less pronounced, with the rate decreasing by 16%, from 77 to 65 per 1,000 women in the same age group. The reduction in unintended pregnancies in developed regions was closely associated with a decrease in abortion rates, while in developing regions, the reduction was more strongly tied to a decrease in unintended births. Between 2010 and 2014, approximately 59% of unintended pregnancies in developed regions and 55% in developing regions resulted in abortion [2].

In Asia, the rate of unintended pregnancies was approximately 54 per 1,000 women aged 15 to 44, resulting in an estimated 53.8 million unintended pregnancies annually during this period [3]. Meanwhile, the global rate of unintended pregnancies in Europe and North America was about 35 per 1,000 women aged 15 to 49. However, in sub-Saharan Africa, the rate was significantly higher, at 91 per 1,000 women [4]. These figures underscore the ongoing disparities in reproductive health across different regions of the world.

Unintended pregnancy exerts a profound influence on various aspects of contemporary life, encompassing social, economic, and cultural dimensions. It is essential to understand the potential repercussions associated with unintended pregnancies. In the United States, nearly half of all unintended pregnancies result in abortion, making it one of the most significant outcomes of unintended pregnancy [5]. Moreover, scholars have explored the potential link between unintended pregnancy and adverse birth outcomes, such as low birthweight, defined as less than 2,500 grams [6]. Low birthweight infants often require extensive resources to support their cognitive, emotional, and relational development, which is crucial for ensuring their overall health and proper development. These needs highlight the long-term impact unintended pregnancies can have, not only on individuals but also on healthcare systems and social services [5]. Limited research exists on the impact of unintended childbearing when. The few studies that have explored adolescent fatherhood and educational attainment have suggested a correlation between teenage fatherhood and an increased likelihood of dropping out of high school. However, these studies have not conclusively determined whether academic difficulties lead to early fatherhood, or if early fatherhood itself contributes to the decision to leave school. This gap in research highlights the need for further investigation into the specific challenges faced by young fathers and how unintended childbearing may affect their educational and life trajectories [5,7].

Prior researchers have found a few common factors associated with unintended pregnancy in Senegal are wealth index, maternal education, intention of contraceptive use [8]. Another critical factor is whether the respondent can refuse physical intimacy. Women who fail to refuse sex with their partners are at higher risk of unintended pregnancy [9]. Another significant predictor is the age of the woman when she had first birth [7]. Moreover, women who made family planning decisions on their own were less likely to have an unintended pregnancy [10]. Women age group tend to influence unintended pregnancy to a great extent [11,12]. These previous studies show that unintended pregnancy is a major concern regarding maternal health which also coincides with many factors. Hence, studying this factor greatly influences future health decisions.

To help reduce unwanted pregnancies, the following suggestions are made: making contraception more widely available; educating people about the significance of motivation, attitudes, and feelings in using contraception and preventing unintended pregnancies; creating and carefully assessing a range of community programs; and promoting research to develop new contraceptives. The new abortion care guideline also suggests simple primary care interventions, such as guaranteeing access to medical abortion pills, making sure that everyone who needs it has access to accurate information, raising the standard of abortion care provided to women and girls, and encouraging task-sharing among a larger range of healthcare professionals [13,14].

This study aligns with the Sustainable Development Goals (SDG) Goal 3: Ensure healthy lives and promote well-being for all at all ages [15]. Which can be essential to achieve SDG goals by creating awareness among women. Traditional approaches to studying unintended pregnancy have primarily relied on descriptive statistics and regression models to identify risk factors (16). While these methods have provided valuable insights, they may fail to capture complex interactions between variables and emerging patterns within large datasets. With the advent of machine learning (ML) techniques, researchers now have the opportunity to leverage advanced analytical tools that can process vast amounts of data and uncover nuanced relationships that may not be immediately apparent through conventional methods [7].

In this study, we apply machine learning techniques to analyze unintended pregnancy in Senegal as unintended pregnancy is highly prevalent there. Between 2015 and 2019 in Senegal, there were approximately 703,000 pregnancies each year. Out of these, 232,000 were unintended pregnancies, and 57,900 of unintended pregnancies resulted in abortion [17]. Moreover, studies had been conducted in Senegal which used classical statistical analysis to understand the correlates of unintended pregnancy [18]. Therefore, machine learning algorithm was utilized to predict unintended pregnancy in this study. By utilizing various machine learning algorithms, we aim to identify key predictors of unintended pregnancy and assess their relative importance. The use of ML allows us to explore intricate associations between demographic, socioeconomic, and behavioral variables while providing more accurate predictions of unintended pregnancy outcomes [19].

This research builds upon existing literature by introducing a data-driven approach to understanding unintended pregnancy in Senegal. Through the application of ML models, we seek to offer more precise and actionable insights that can inform public health policies and programs aimed at reducing unintended pregnancies in the region. Ultimately, our goal is to contribute to improving reproductive health outcomes and empowering women in Senegal to make informed choices about their reproductive lives.

## Methods

### Data Source and Population

This research utilized nationally representative secondary data from the 2023 Senegal Demographic and Health Survey which was conducted from February 2023 to August 2023. The Demographic Health Survey Authority employed a two-stage stratified sampling method. The data was collected from 14 regions namely: Dakar, Ziguinchor, Diourbel, Saint-Louis, Tambacounda, Kaolack, Thies, Louga, Fatick, Kolda, Matam, Kaffrine, Kedougou and Sedhiou. Extensive data were collected from 8423 households, 16583 female respondents who were aged between 15 to 49, and 6321 male respondents who were aged between 15 to 59, covering a wide range of topics. These included adult and childhood morbidity and mortality, awareness and attitudes toward HIV/AIDS, fertility and fertility preferences, marriage, knowledge and use of family planning methods, and various aspects of reproductive health, among other significant public health concerns. This study was used conducted on the women who were pregnant at the time of conducting the survey and the number of respondents was 1065 persons.

### Study Variables

The dependent variable was whether the respondent was pregnant at the time of the survey which had three (3) type of responses: Then, Later and Not at all. To evaluate respondent’s pregnancy intentions, the responses were recoded as:

1. ‘Then’ for ‘Intended’ which was codes as zero (0).
2. ‘Later’ and ‘Not at all’ for ‘Unintended’ which as coded as one (1)

The explanatory variables that were used in this study are respondent’s age group (15 to 19, 20 to 24, 25 to 29, 30 to 49), region (Dakar, Ziguinchor, Diourbel, Saint-Louis, Tambacounda, Kaolack, Thies, Louga, Fatick, Kolda, Matam, Kaffrine, Kedougou, Sedhiou), residence type (urban, rural), respondent’s education level (No education, Primary education, Secondary and higher), wealth index (poor, middle, rich), number of living children (0, 1-2, 3+), total birth (0, 1-2, 3+), intention of contraceptive use (Intended to use, Unintended to use), currently residing with husband/partner (living with partner, staying elsewhere), Respondent can refuse sex (Yes, No), age at first cohabitation (<18, ≥18), Ideal number of children (<6, ≥6), Husband/partner’s education level (No education, Primary education, Secondary and higher), Husband/partner’s occupation (Not, employed, Employed, Don’t know) and Respondent occupation (Not, employed, Employed).

### Data Processing and Analysis

The number of missing values were less than 5% (<5%) and hence the observations were deleted for those missing values. The final dataset had reduced observations of 885 respondents. A basic descriptive analysis followed by a bivariate analysis was performed in the study. The descriptive analysis was used to outline the frequency and percentage distribution of the data. We then employed bivariate analysis to explore the association and correlation. Six different supervised machine learning algorithms were used to predict the outcome variable and assessed their performance using various model evaluation metrics. The machine learning models are namely Logistic Regression (LR), Random Forest (RF), K-Nearest Neighbors (KNN), Support Vector Machine (SVM), Naïve Bayes (NB) and Extra Tree Classifier (ETC). Data processing was conducted in SPSS version 23 and Python. **Figure 1** displays the theoretical framework of the entire analysis procedure.

**Figure 1:**
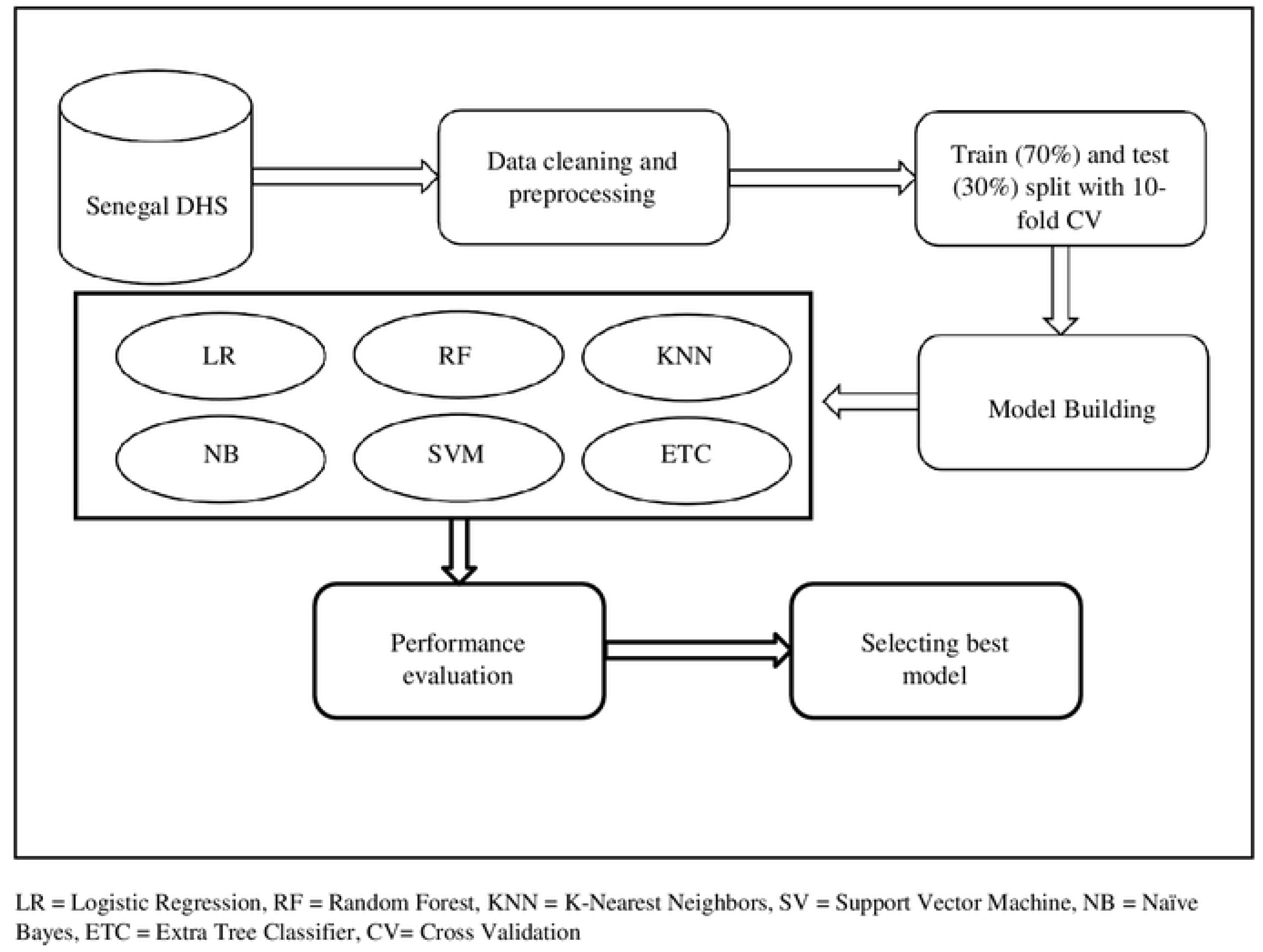
Theoretical Framework of Predicting Unintended Pregnancy using ML

#### Logistic Regression (LR)

Specifically created for “classification” tasks, logistic regression (LR) is a “statistical learning” approach that falls under the “supervised” machine learning (ML) category. To estimate the parameters of interest, the maximum likelihood estimation approach is utilized [20]. The logistic regression equation models the probability p of a binary outcome (e.g., 0 or 1) as a function of predictor variables X. It is represented as:

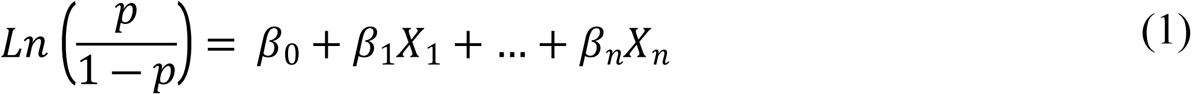

Where, p is the probability of the dependent variable (unintended pregnancy), ß_0,_ ß_1,…,_ ß_n_ are the coefficients of predictor variables and X_1_,…, X_n_ are the predictor variables.

#### Random Forest (RF)

One of the well-known supervised learning methods is the Random Forest algorithm. This machine learning technique can be applied to both classification and regression problems. It is based on the concept of ensemble learning, which involves combining multiple classifiers to address complex issues and improve the model’s performance [21].

#### K-Nearest Neighbors (KNN)

A non-parametric supervised learning classifier, the k-nearest neighbors (KNN) technique uses the proximity of data points to classify or predict which category a given data point belongs to [22]. KNN finds the distance between the query point X_q_ and all other points in the dataset. The most common distance metric is the Euclidean distance which is a measure of the true straight-line distance between two points in Euclidean space [23]. This distance is measured given by the formula:

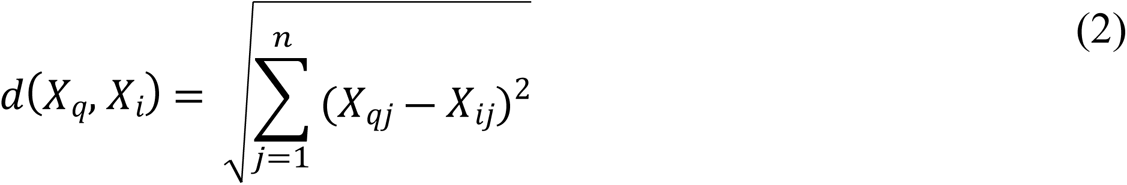

Where d(X_q_, X_i_) is the Euclidean distance between the query point X_q_ and the i-th point X_i_, n is the number of features (dimensions) and X_qj_ and X_ij_ are the values of the j-th feature for the query point and the i-th data point, respectively.

#### Support Vector Machine (SVM)

Strong machine learning algorithms like Support Vector Machine (SVM) are utilized for tasks including regression, outlier identification, and linear or nonlinear classification. Because SVMs can handle high-dimensional data and nonlinear relationships, they are versatile and effective in a wide range of applications [24].

#### Naïve Bayes (NB)

A group of classification algorithms based on Bayes’ Theorem are known as Naive Bayes classifiers. It is not a single algorithm, but rather a collection of algorithms that are united by a basic principle: each pair of features being classed stands alone. Bayes’ Theorem calculates the likelihood of an event happening based on the probability of a related event that has already taken place [25]. Mathematically, it is expressed as:

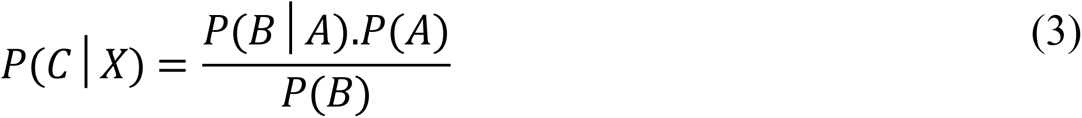

Where P(C|X) is the posterior probability of class (C, target) given predictor (X, attributes). P(C) is the prior probability of class, P(X|C) is the likelihood which is the probability of the predictor given class, P(X) is the prior probability of the predictor.

#### Extra Tree Classifier (ETC)

Extra Tree Classifier is an ensemble machine learning technique that trains multiple decision trees and then combines their output to produce a forecast which can be used to build classification model or regression models [26]. This can be expressed as:

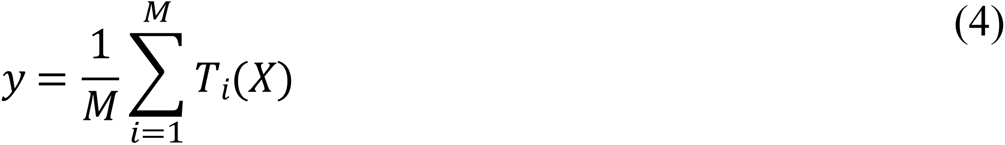

Where y is the predicted class for input X, T_i_(X) is the prediction made by the i-th tree for input X and M is the total number of trees in the ensemble.

### Feature Selection

Ranking and prioritizing the most significant predictors in the dataset is the aim of feature selection. The information gain values for each of the chosen variables are computed to determine this. The amount of knowledge a feature in a classification problem offers about a class is measured as information gain. Decision tree algorithms is used to find the most informative features for data splitting. When predicting the class labels of newly discovered data instances, features with a high information gain hold greater significance. The information gain, which varies from 0 (no gain) to 1 (highest gain), is computed using the entropy of the class distribution both before and after the split [27].

We have selected the top 8 variables according to information gain value for machine learning algorithms. The ranked variables are displayed in **Figure 2**. The features that were selected according to information gain value for machine learning algorithms are total birth, currently residing with husband/partner, respondent’s education level, number of living children, Husband/partner’s occupation, residence type, Respondent can refuse sex and intention of contraceptive use.

**Figure 2:**
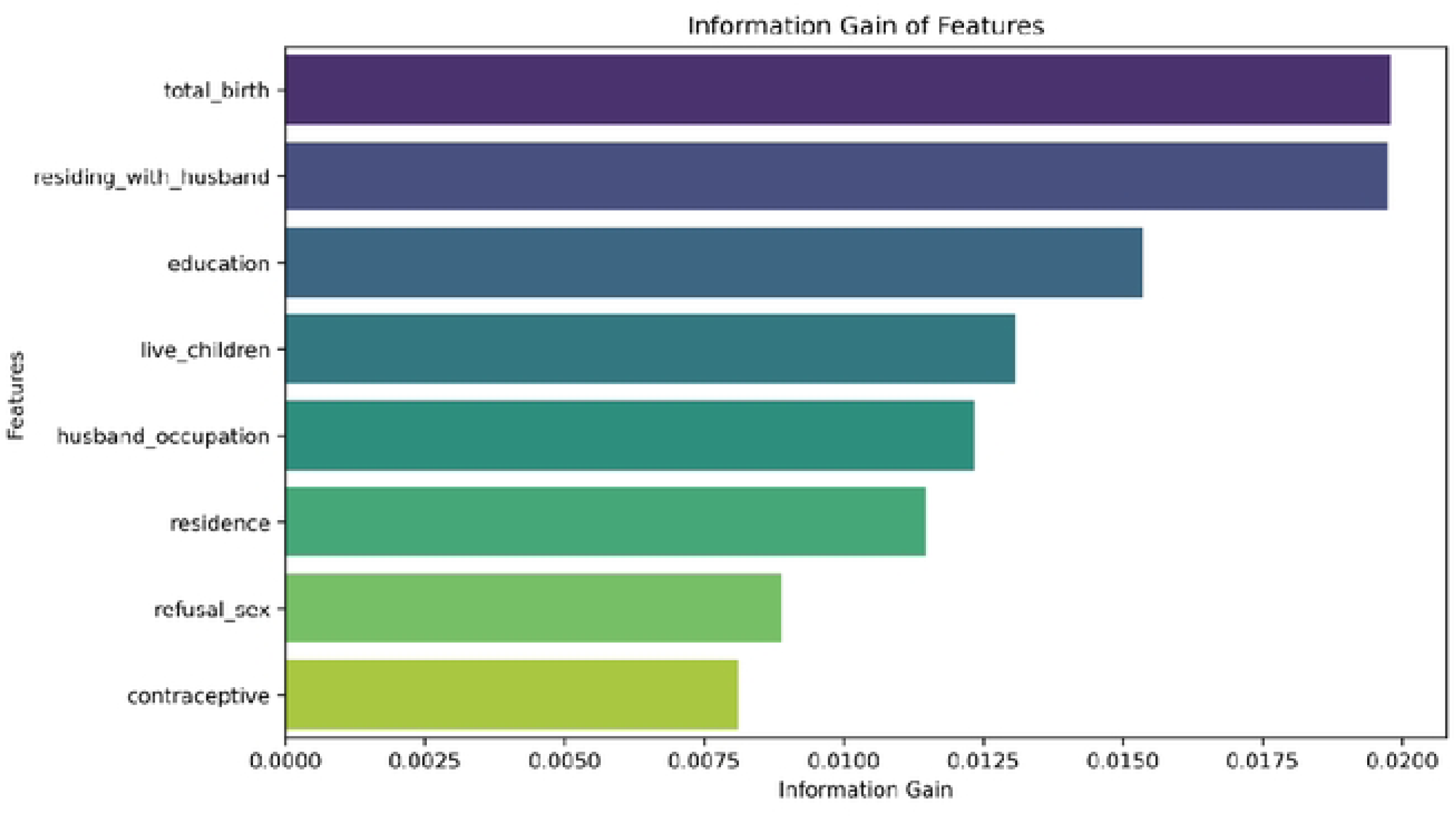
Variables Ranked according to their Information Gain

### Data split

The dataset is randomly split into two sections for machine learning approaches: a training dataset, which is used to train the model, and a test dataset, which is used to predict the response variable and determine if the predicted results match the actual results. In this study, 70% of the total dataset was taken as training dataset and the remaining 30% was taken as testing dataset. On the training set, we employed 10-fold repeated cross-validation and assessed the performance on the testing dataset.

### Handling Imbalance Dataset

The dataset which had been used for the purpose of the study was heavily imbalanced. There were 885 respondents on the final dataset in which 763 (86.2%) respondents had intended pregnancy and the remaining 122 (13.8%) respondents had unintended pregnancy. Because an imbalanced dataset tends to be skewed towards the majority class and results in low performance on the minority class, it was necessary to balance the classes. For the minority class, this may lead to great accuracy but poor precision and recall [28].

In this work, the Synthetic Minority Oversampling Technique (SMOTE) was utilized to maintain a balance between the majority and minority classes. Through the use of oversampling imbalanced datasets, SMOTE is a pre-processing technique for learning algorithms that effectively addresses class imbalance. It creates a new sample by randomly layering a few samples linearly on top of their neighbors [29]. **Figure 3** represents the balancing of classes before and after applying SMOTE.

**Figure 3:**
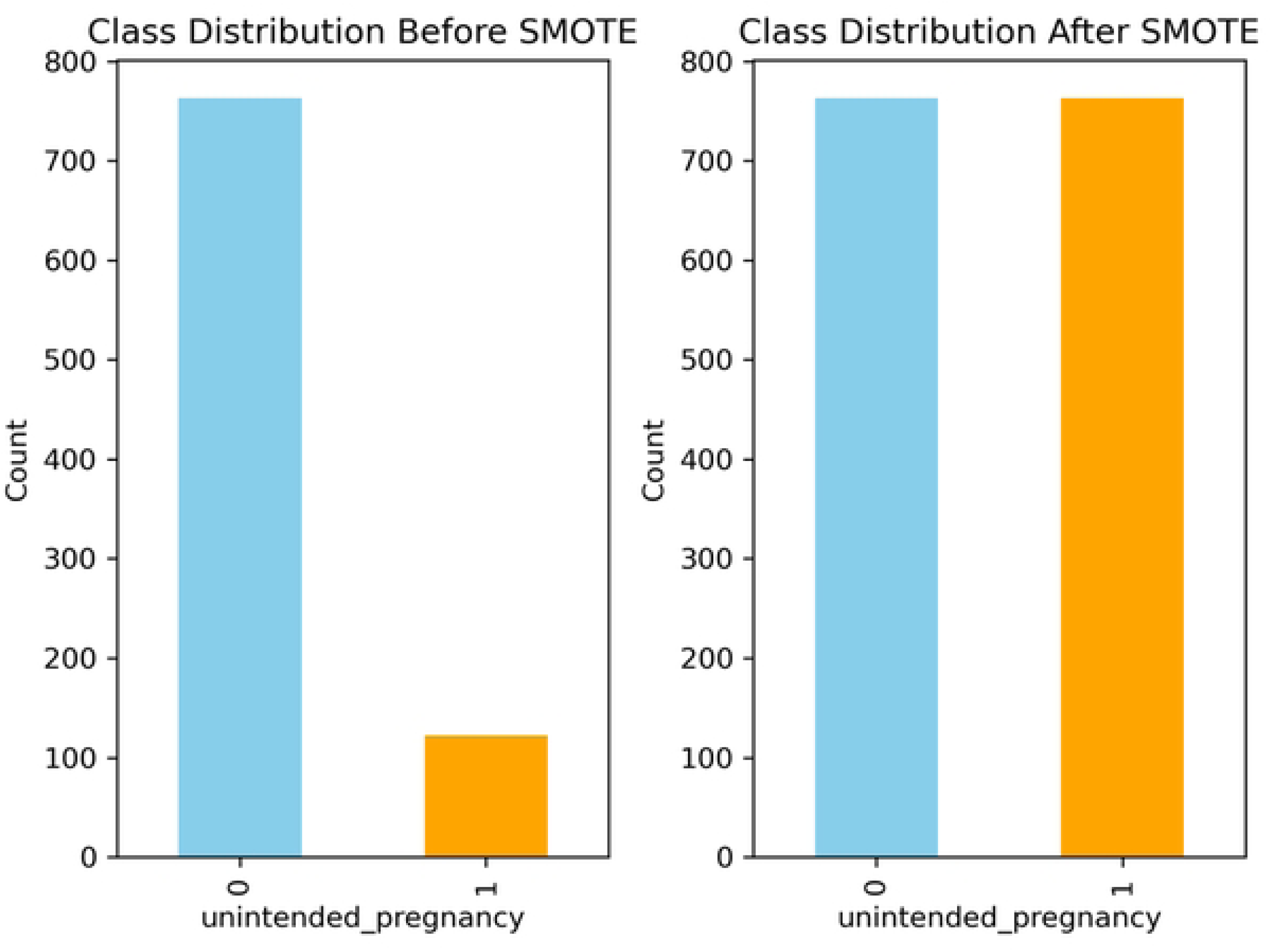
Class Distribution Before and after SMOTE

### Performance Evaluation for Machine Learning Algorithms

Every model’s performance is evaluated and compared to other models after training. The performance of the prediction models was evaluated using the accuracy, precision, recall, specificity, F1-score, and area under the curve (AUC-Receiver Operating Characteristic) were applied in this investigation to assess the performance of the model.

#### Accuracy

Accuracy is an assessment parameter used in machine learning that measures how well a model predicts the future overall. It shows the proportion of accurately predicted cases, true positives and true negatives, to all instances in the dataset [30].

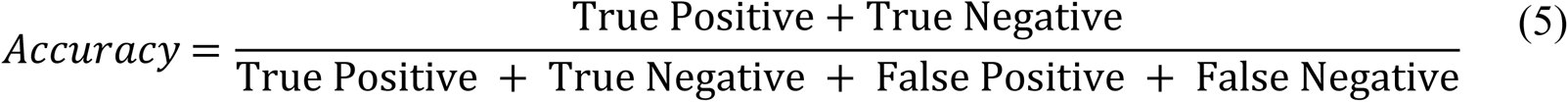

#### Precision

The precision is the percentage of all positive classifications made by the model that are true positives [31]. It is mathematically defined as:

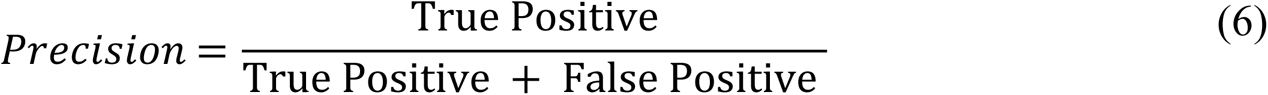

#### Recall

Recall is determined by dividing the total number of Positive samples by the number of Positive samples that were accurately categorized as Positive. The recall gauges how well the model can identify positive samples. Positive samples are found in greater numbers the higher the recall [32].

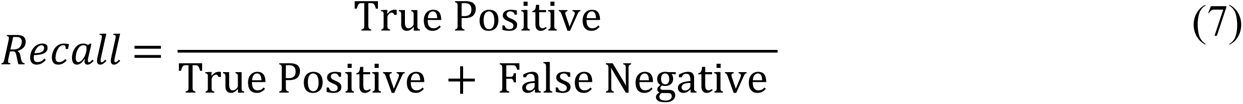

#### F1-Score

An alternative machine learning evaluation statistic called the F1 score elaborates on a model’s performance within a class, as opposed to evaluating the model’s overall performance based on accuracy. The F1 score integrates a model’s precision and recall ratings, two conflicting criteria [33].

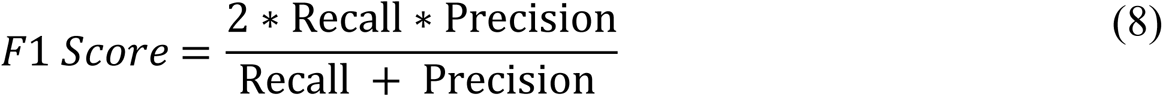

#### Specificity

The ability of an algorithm or model to accurately anticipate a true negative for every category that exists is measured by machine learning specificity. In academic settings, the term “true negative rate” is frequently used [34]; its formal calculation is:

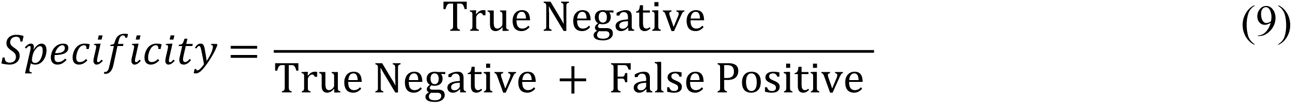

#### AUC Value

The area under the Receiver Operating Characteristic curve is represented by the AUC curve, which stands for Area Under the Curve. It measures the binary classification model’s overall effectiveness. The area will always lie between 0 and 1, since both TPR and FPR vary from 0 to 1. A higher AUC value indicates better model performance [35].

## Results

### Sociodemographic Characteristics

Table 1 provides an overview of the background characteristics of participants with unintended pregnancies. The highest percentage of respondents were from Matam (12.9%), while the lowest percentage were from Ziguinchor (2.6%). A significant portion of the participants (35.1%) fell within the 30-49 age group, which encompasses a broader age range. Additionally, 27.5% of participants were aged between 20-24 years. The majority of respondents resided in rural areas (67.1%).

**Table 1:**
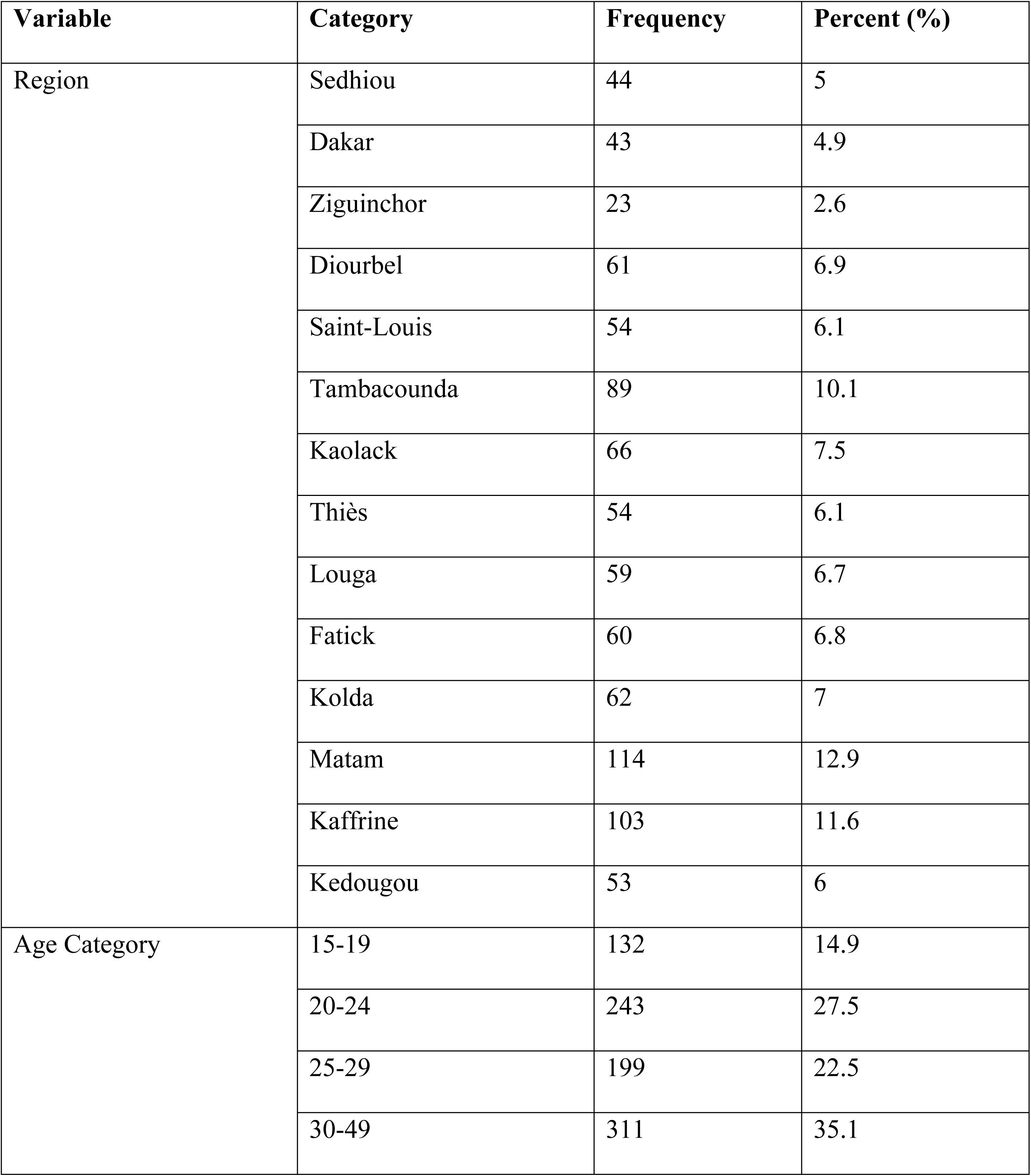
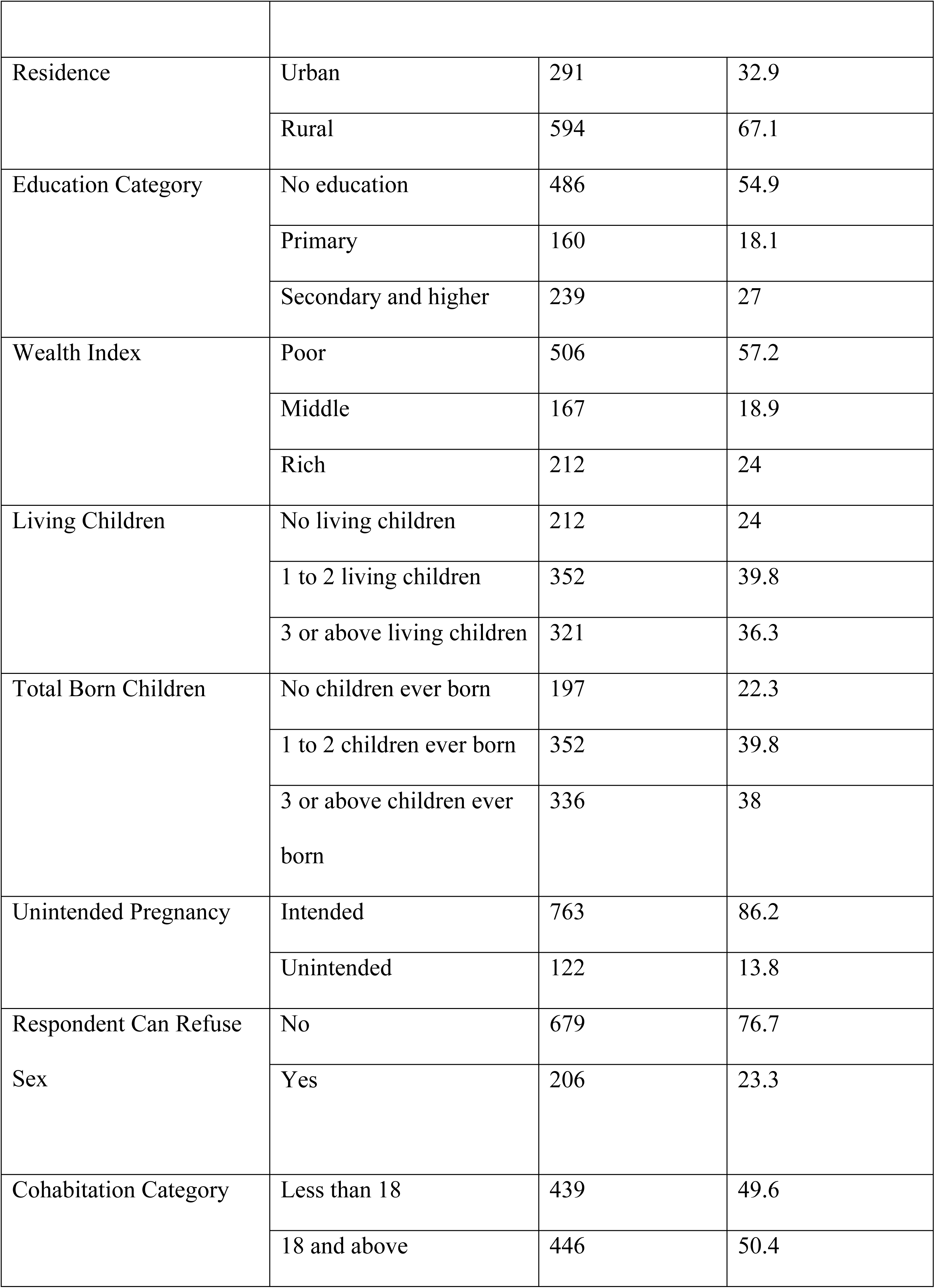
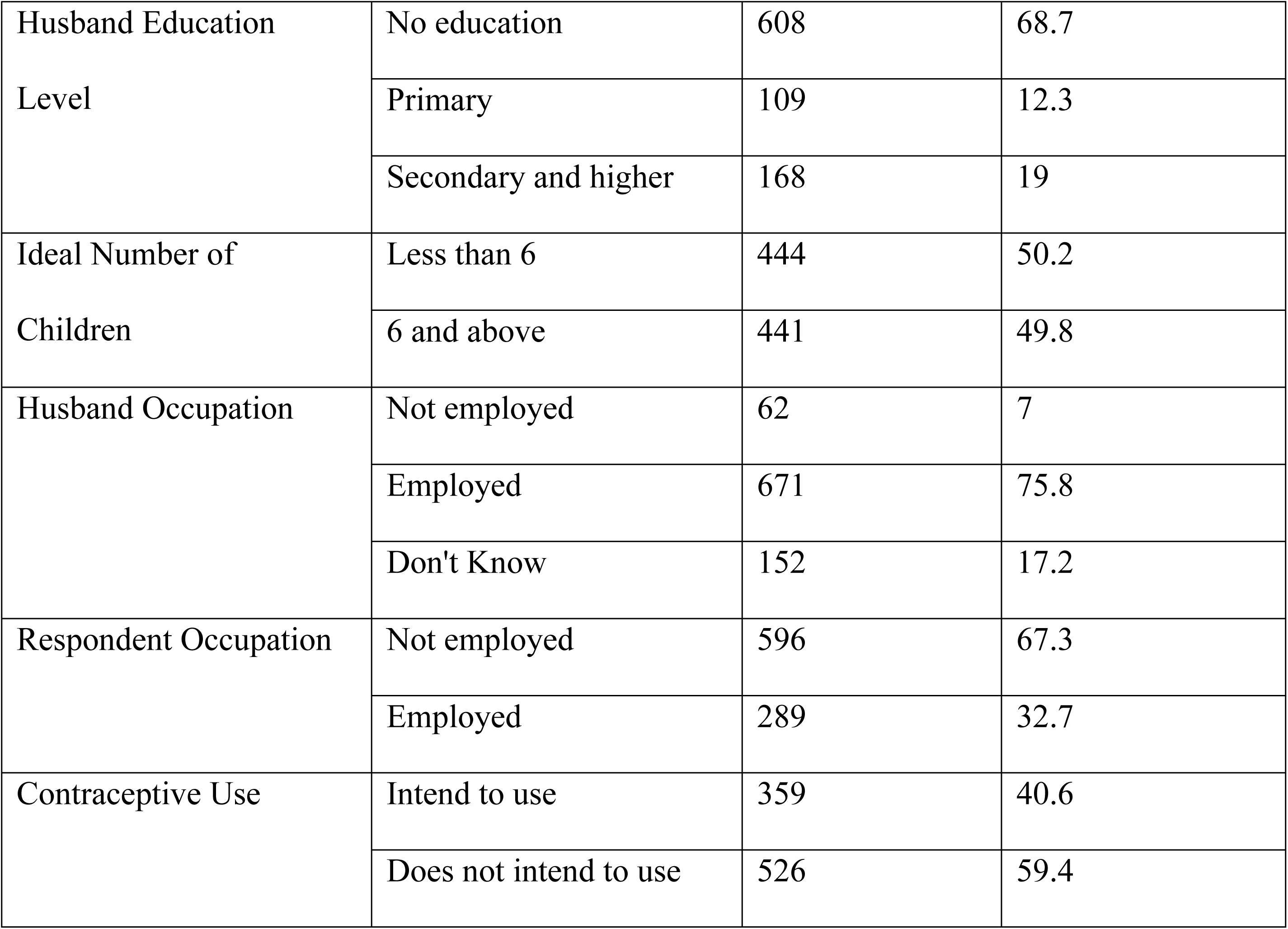
Background characteristics of the study participants.

| Variable | Category | Frequency | Percent (%) |
| --- | --- | --- | --- |
| Region | Sedhiou | 44 | 5 |
|  | Dakar | 43 | 4.9 |
|  | Ziguinchor | 23 | 2.6 |
|  | Diourbel | 61 | 6.9 |
|  | Saint-Louis | 54 | 6.1 |
|  | Tambacounda | 89 | 10.1 |
|  | Kaolack | 66 | 7.5 |
|  | Thiès | 54 | 6.1 |
|  | Louga | 59 | 6.7 |
|  | Fatick | 60 | 6.8 |
|  | Kolda | 62 | 7 |
|  | Matam | 114 | 12.9 |
|  | Kaffrine | 103 | 11.6 |
|  | Kedougou | 53 | 6 |
| Age Category | 15-19 | 132 | 14.9 |
|  | 20-24 | 243 | 27.5 |
|  | 25-29 | 199 | 22.5 |
|  | 30-49 | 311 | 35.1 |
| Residence | Urban | 291 | 32.9 |
|  | Rural | 594 | 67.1 |
| Education Category | No education | 486 | 54.9 |
|  | Primary | 160 | 18.1 |
|  | Secondary and higher | 239 | 27 |
| Wealth Index | Poor | 506 | 57.2 |
|  | Middle | 167 | 18.9 |
|  | Rich | 212 | 24 |
| Living Children | No living children | 212 | 24 |
|  | 1 to 2 living children | 352 | 39.8 |
|  | 3 or above living children | 321 | 36.3 |
| Total Born Children | No children ever born | 197 | 22.3 |
|  | 1 to 2 children ever born | 352 | 39.8 |
|  | 3 or above children ever born | 336 | 38 |
| Unintended Pregnancy | Intended | 763 | 86.2 |
|  | Unintended | 122 | 13.8 |
| Respondent Can Refuse Sex | No | 679 | 76.7 |
|  | Yes | 206 | 23.3 |
| Cohabitation Category | Less than 18 | 439 | 49.6 |
|  | 18 and above | 446 | 50.4 |
| Husband Education Level | No education | 608 | 68.7 |
|  | Primary | 109 | 12.3 |
|  | Secondary and higher | 168 | 19 |
| Ideal Number of Children | Less than 6 | 444 | 50.2 |
|  | 6 and above | 441 | 49.8 |
| Husband Occupation | Not employed | 62 | 7 |
|  | Employed | 671 | 75.8 |
|  | Don't Know | 152 | 17.2 |
| Respondent Occupation | Not employed | 596 | 67.3 |
|  | Employed | 289 | 32.7 |
| Contraceptive Use | Intend to use | 359 | 40.6 |
|  | Does not intend to use | 526 | 59.4 |

Regarding education, most participants had no formal education (54.9%). In terms of socioeconomic status, a majority of respondents were categorized as poor (57.2%), while middle-class respondents represented the smallest proportion. Notably, the incidence of unintended pregnancy was relatively low at 13.8%. In most cases (76.7%), respondents reported that they were unable to refuse sex. It is also concerning that the majority of respondents’ husbands lacked formal education, with 68.7% having received no education. Moreover, 75.8% of the husbands were employed whereas only 32.7% of the respondents were employed. Majority of the respondents (59.4%) does not intend to use contraceptive.

Table 2 presents the prevalence of unintended pregnancy in relation to various background characteristics and highlights the significance of these associations. Region significantly influences the likelihood of pregnancy intention, with Ziguinchor exhibiting the highest percentage of intended pregnancies (95.7%, p-value = .001). However, age categories do not show a significant impact on unintended pregnancy rates. The residence variable is significantly associated with unintended pregnancy (p-value = .059). Other factors, such as education level, the respondent’s ability to refuse sex, cohabitation age, husband’s education level, ideal number of children, and husband’s occupation, do not significantly influence unintended pregnancy.

**Table 2:**
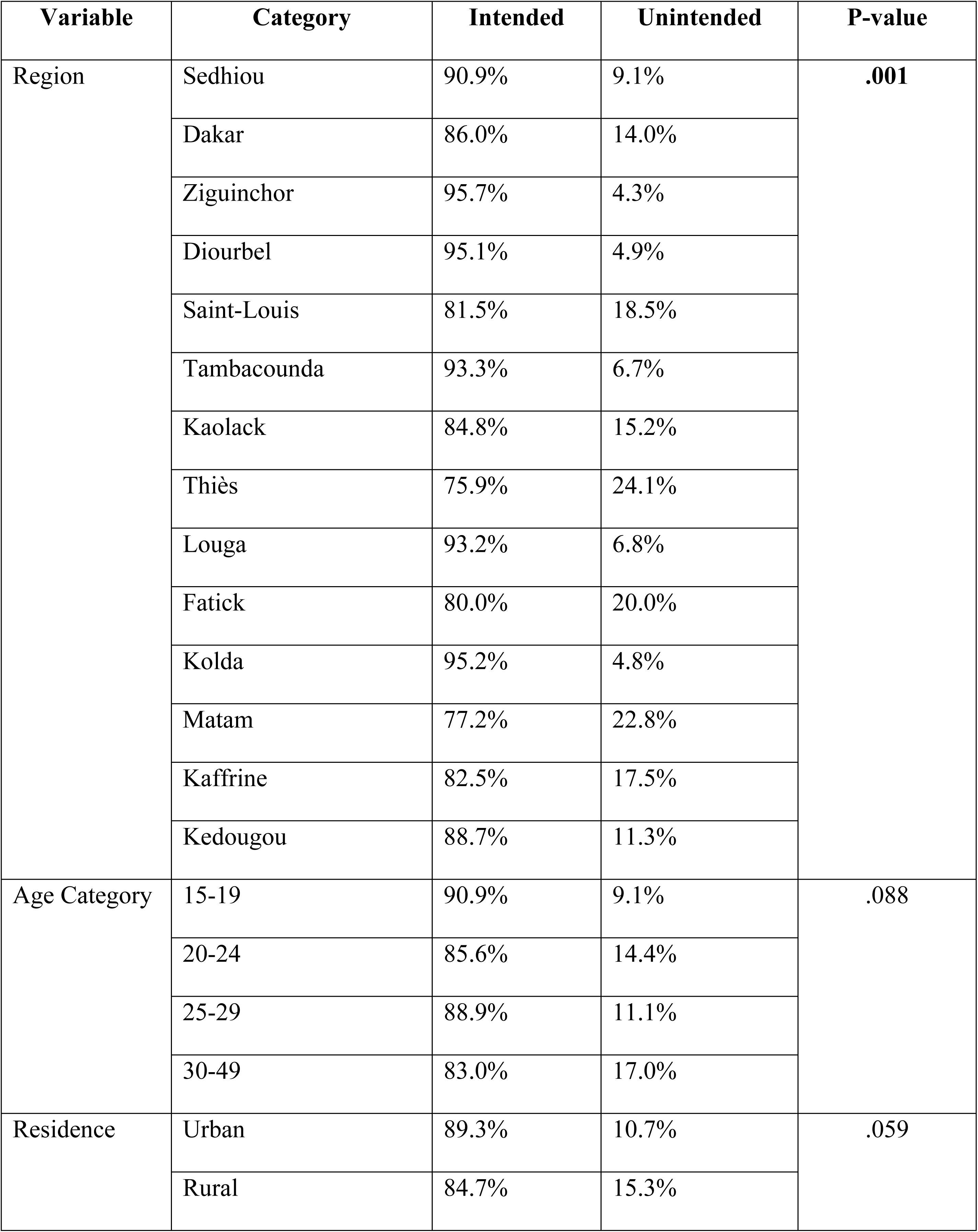
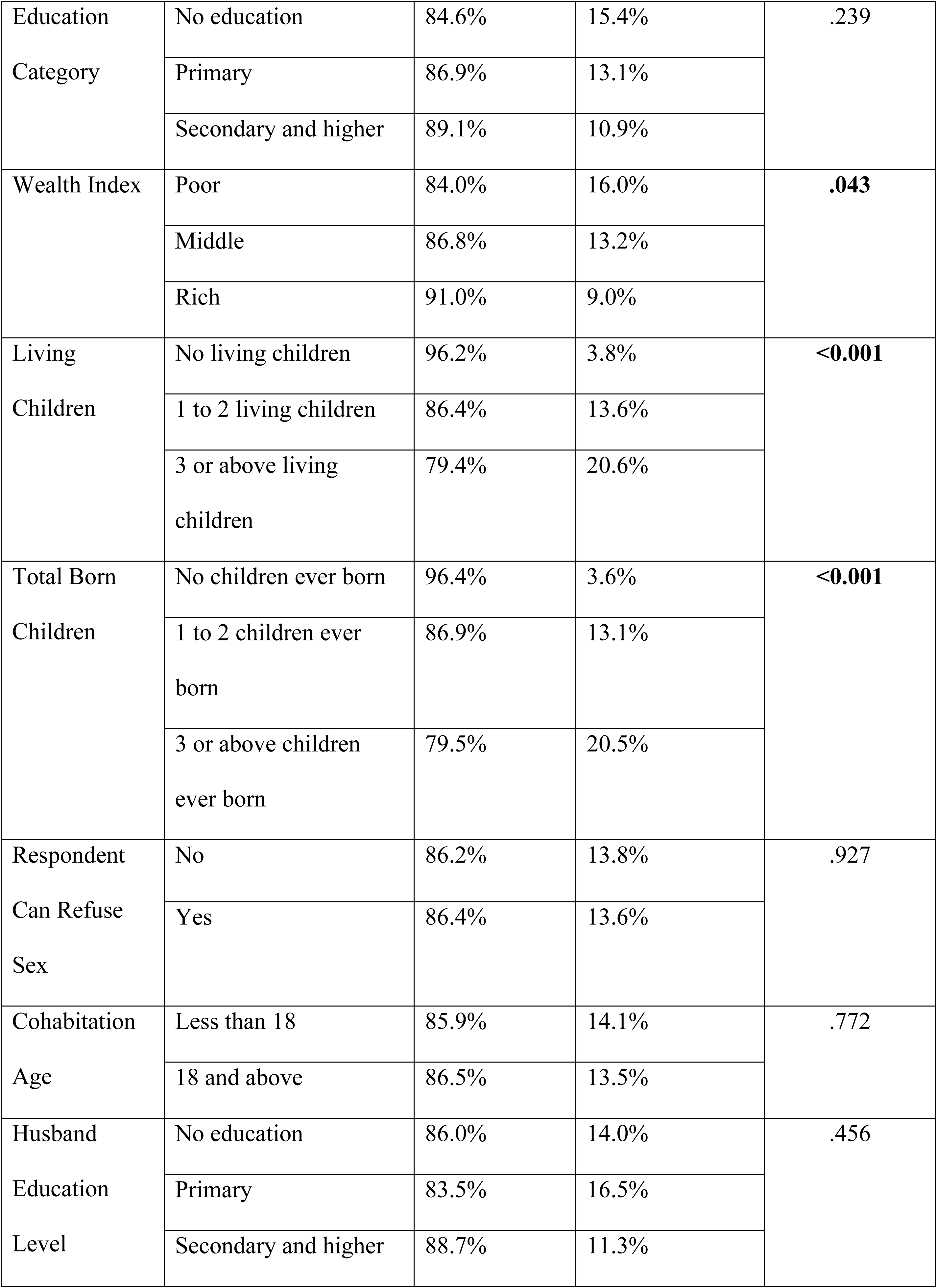
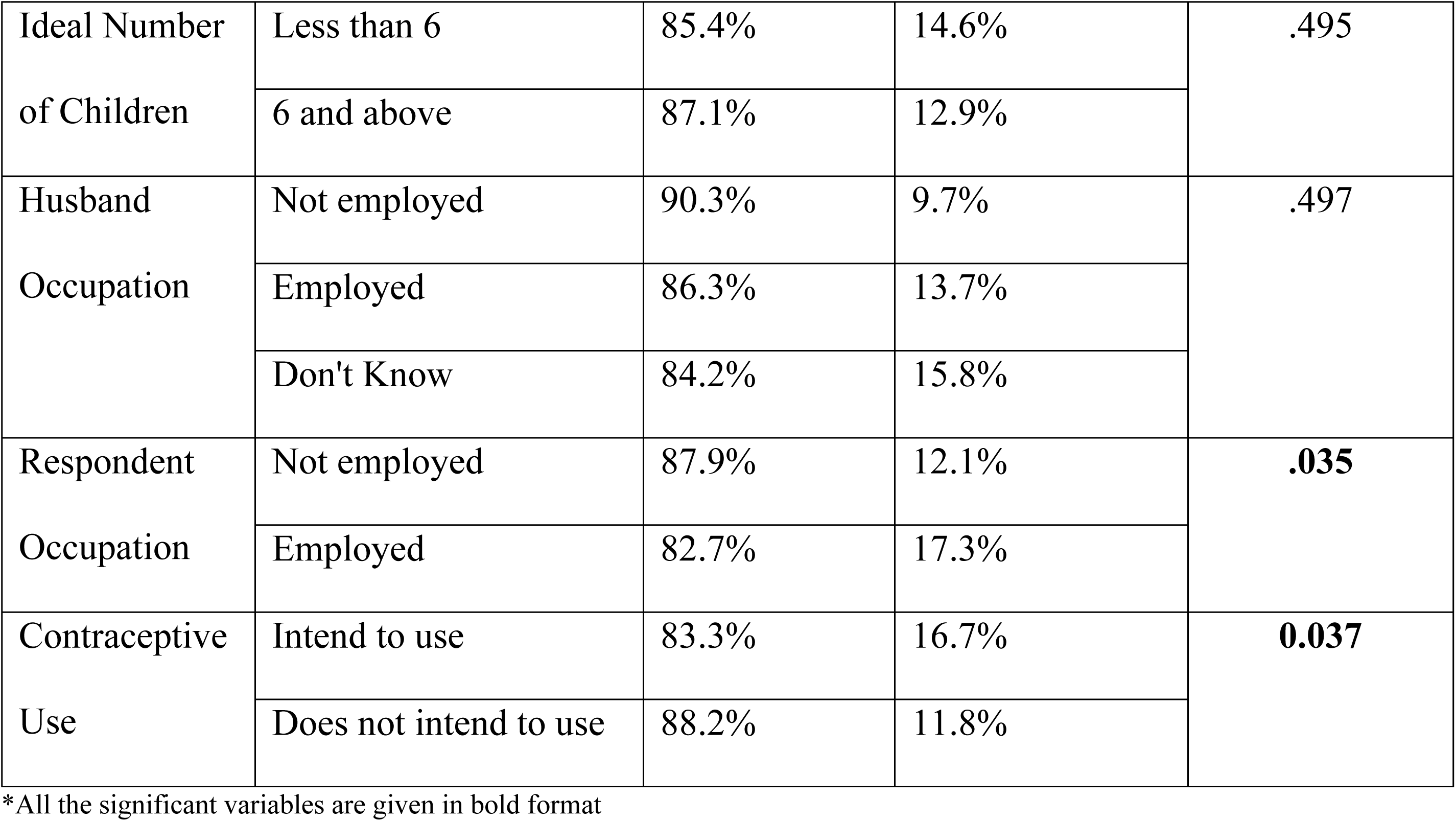
Percentage distribution and association between selected covariates and women’s pregnancy intentions in Bangladesh.

Additionally, the data reveal that women with a higher economic status (rich) have a lower unintended pregnancy rate (91.0%). Women with no living children show the lowest rate of unintended pregnancy (96.2%, p-value < .001). Furthermore, non-employed women (87.9%, p-value = .035) and those who do not intend to use contraceptives (88.2%, p-value = .037) also exhibit lower rates of unintended pregnancy.

### Implementation of Machine Learning

In this study, the data were divided into 70:30 ratio were 70% of the data was the training dataset and the remaining (30%) was the testing dataset. In order to categorize the women in the test dataset into intended and unexpected pregnancies, six distinct machine learning algorithms were employed. AUC value, sensitivity, specificity, accuracy, precision and F1 score were utilized as performance metrics to compare the prediction performance of these algorithms. Table 3 and Figure 3 display the prediction output together with performance metrics for each machine learning algorithm.

**Table 3:** Performance of Machine Learning Algorithms.

|  | LR | RF | KNN | SVM | NB | ETC |
| --- | --- | --- | --- | --- | --- | --- |
| Accuracy | 0.7064 | 0.7339 | 0.5727 | 0.6041 | 0.6998 | 0.7392 |
| (95% CI) | (0.67, 0.73) | (0.72, 0.74) | (0.55, 0.59) | (0.57, 0.63) | (0.67, (0.72) | (0.72, 0.75) |
| Precision | 0.6901 | 0.6939 | 0.7244 | 0.5780 | 0.6816 | 0.7013 |
| Recall | 0.7562 | 0.8388 | 0.2308 | 0.7695 | 0.7509 | 0.8349 |
| F1-Score | 0.7204 | 0.7589 | 0.3478 | 0.6568 | 0.7141 | 0.7618 |
| Specificity | 0.6565 | 0.6290 | 0.9147 | 0.4392 | 0.6487 | 0.6435 |
| AUC Value | 0.7646 | 0.8045 | 0.6787 | 0.6453 | 0.7557 | 0.7920 |

LR = Logistic Regression, RF = Random Forest, KNN = K-Nearest Neighbors, SV = Support Vector Machine, NB = Naïve Bayes,
ETC = Extra Tree Classifier, CI = Confidence Interval

In Table 3, the accuracy for Random Forest (RF) classifier and Extra Tree Classifier (ETC) is 73.39% and 73.92% which means these two models are 73% correct for predicting unintended pregnancy. Logistic Regression (LR) classifier has an accuracy of 70.64% and the remaining three machine learning algorithms (K-Nearest Neighbors, Support Vector Machine and Naïve Bayes) have accuracy below 70%. In case of precision, K-Nearest Neighbors have the highest precision (72.44%) and Extra Tree Classifier has 70.13% precision. The remaining four algorithm have precision less than 70%. The recall value for Random Forest and Extra Tree Classifier are 83.88% and 83.49% respectively which are higher than the remaining four algorithms. Similarly, Random Forest and Extra Tree Classifier have higher F1 Scores, 75.89% and 76.18% respectively, and the remaining classifiers have less than 75% F1 Scores. K-Nearest Neighbors has the highest specificity value, 91.47%, which is highest among all six machine learning algorithms. On the other hand, Random Forest algorithm has the highest AUC value which is 80.45%. The remaining five machine learning algorithm has lower than 80% AUC value.

Among all six machine learning algorithms, Random Forest and Extra Tree Classifier have the highest accuracy. According to ROC (Receiver Operating Characteristic) Curve and AUC value displayed in **Figure 4**, Random Forest algorithm has the highest AUC value and hence it was selected as the best algorithm for predicting unintended pregnancy in Senegal.

**Figure 4:**
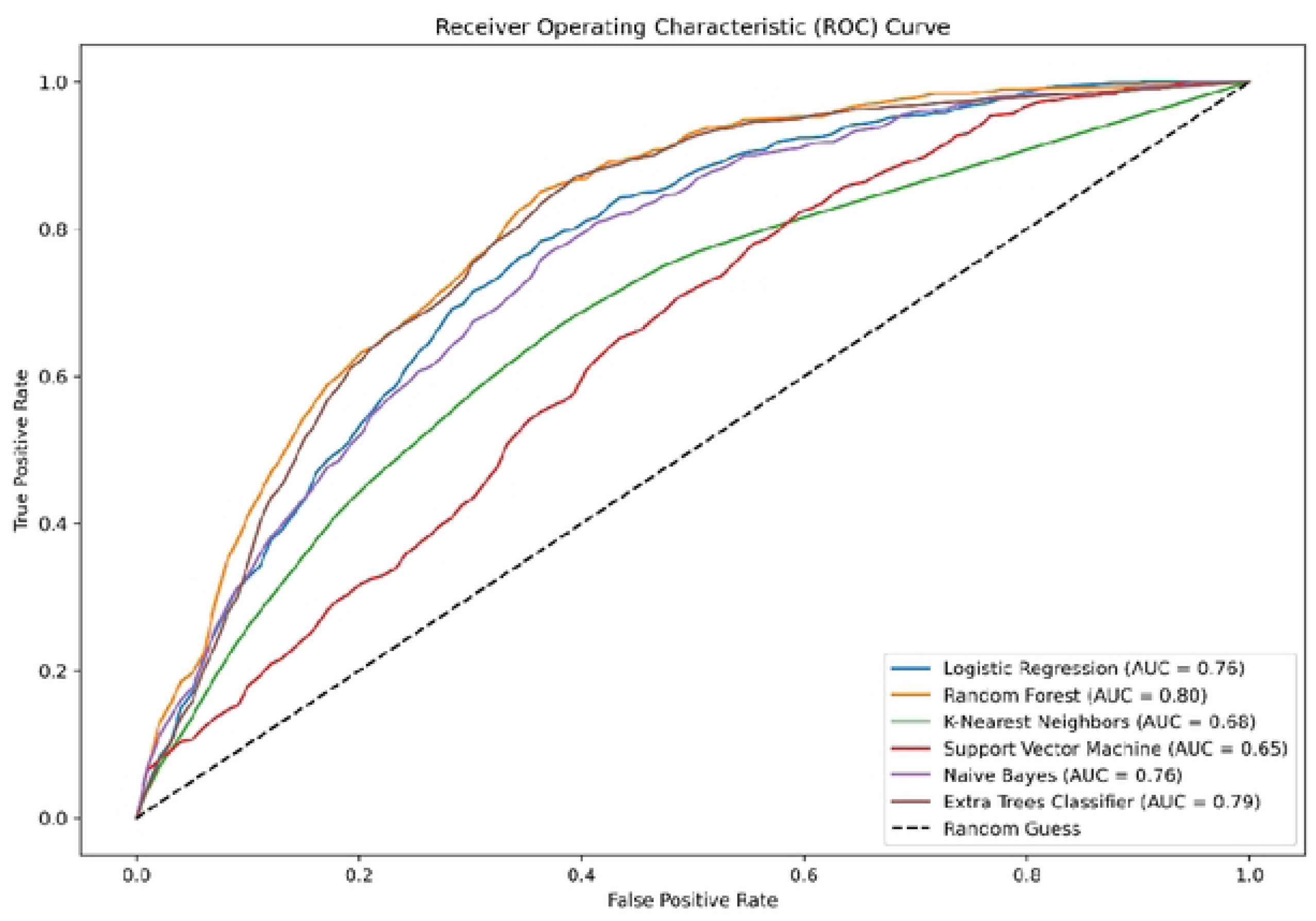
ROC Curve for Six Machine Learning Algorithms

### Tuning Random Forest Classifier with Grid Search CV

This study used hyperparameter tuning to compare the optimal model with the default hyperparameter tuning once the best model was chosen. **Figure 5** demonstrates that the default hyperparameter tuning performed better than the hyperparameter tuning with the Random Forest Classifier. As per the findings, the Random Forest with adjusted hyperparameters performed less effectively than the Random Forest model with the default hyperparameter.

**Figure 5:**
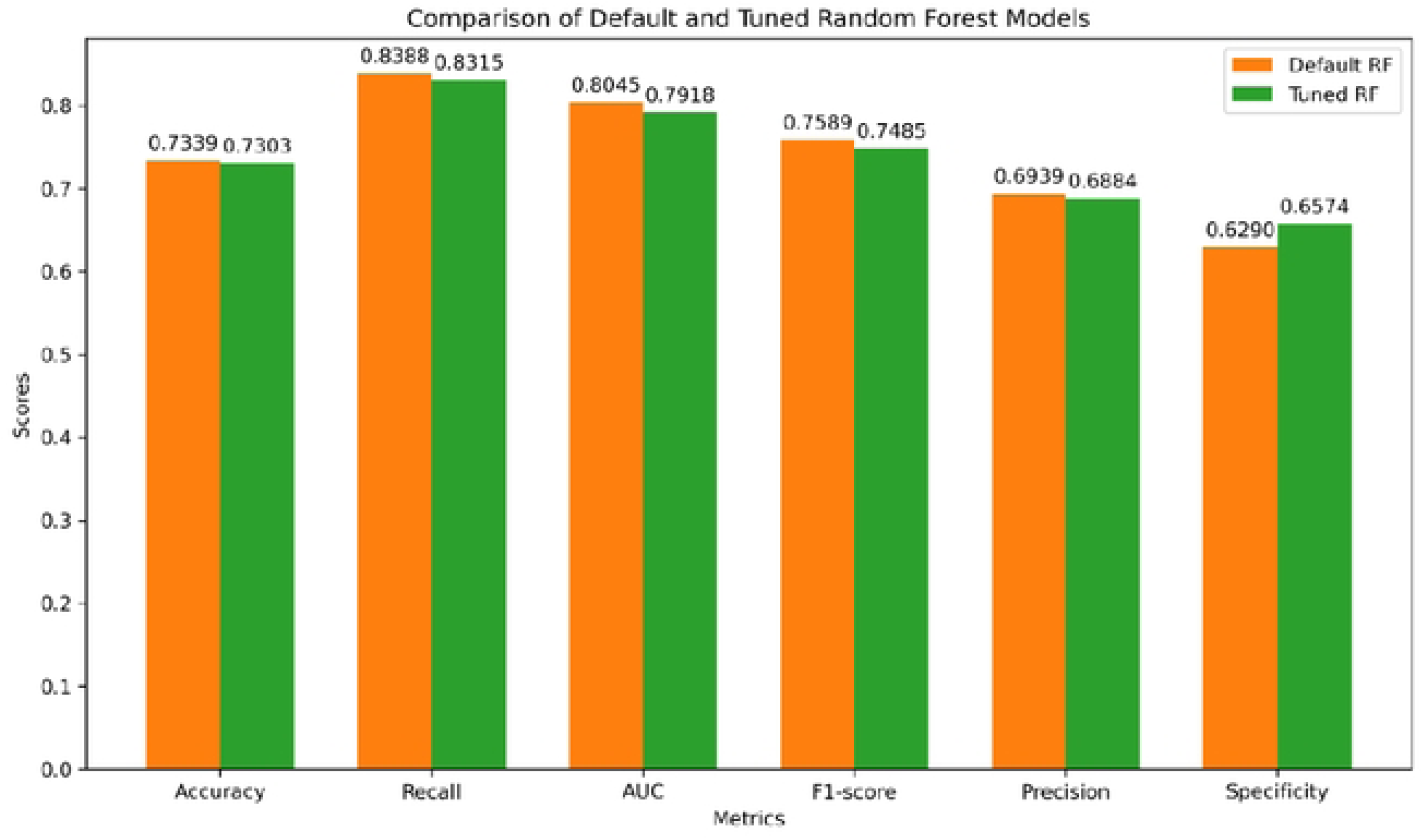
Comparison of Tuned and Default Hyperparameters of Random Forest Algorithm

From Figure 4, it is seen that the accuracy, recall, AUC, F1 score and precision are higher for the default Random Forest model than that of the tuned Random Forest model. However, in case of specificity, the default Random Forest model has 62.90% specificity which is lower than that of tuned Random Forest model (65.74%). The Random Forest model with default hyperparameter tuning obtained the highest AUC score and accuracy, indicating that it correctly detected unintended pregnancy. Hence, the default parameter Random Forest model was selected as the best model for predicting unintended pregnancy in Senegal.

## Discussion

There have studies related to unintended pregnancy in Senegal. However, to our knowledge, this is the first study in Senegal which have utilized machine learning algorithms to predict unintended pregnancy and also identify the potential factors. Machine learning algorithms are proven to be more efficient and accurate for prediction performance than classical statistical analysis [36]. The purpose of this study was to identify potential risk factors for unintended pregnancy and making predictions using six different machine learning algorithms among Senegalese women. Six well-known machine learning algorithms which were applied to meet the research purpose are Logistic Regression, Random Forest, K-Nearest Neighbor, Support Vector Machine, Naive Bayes, and Extra Tree Classifier and these algorithms are widely renowned for their classification tasks. The Senegal Demographic and Health Survey 2023 data was used to conduct the entire analysis and research.

The prediction performance of these six machine learning methods is compared based on the curve value area or AUC value and accuracy. The dataset was divided into training dataset (70%) and testing dataset (30%). The classifiers were trained on the training data using a stratified 10-fold cross-validation approach. According to accuracy, the best machine learning algorithm was Random Forest and Extra Tree Classifier as they both had almost similar accuracy, 73.39% and 73.92%, Extra Tree Classifier having a slightly higher accuracy than Random Forest. On the contrary, Random Forest have a higher AUC value than Extra Tree Classifier, 80.45% and 79.20% which clearly made Random Forest the best predictive model among all six machine learning algorithms.

The results of our study are similar to that of another study conducted in Missouri where Random Forest algorithm had the highest AUC value and was selected as the best model for predicting unintended pregnancy [37]. On the contrary, Extra Tree Classifier was selected as the best algorithm for predicting unintended pregnancy in Ethiopia as it had higher performance evaluation metrics than other models [19]. A similar study conducted among Bangladeshi women identified Elastic Net Regression (ENR) as the best model for predicting unintended pregnancy in Bangladesh [7]. In a different setting, a study conducted on predicting pregnancy outcome following IVF (In Vitro Fertilization) treatment suggested that Support Vector Machine (SVM), Random Forest (RF) and Multilayer Perceptron (MLP) yielded better results than all other machine learning algorithms [38]. Random Forest performed best in another study regarding the prediction of adverse pregnancy outcomes in pregnant women with systemic lupus erythematosus, A chronic autoimmune inflammatory disease that involves multiple organs and predominantly affects women of reproductive age in China [39]. Moreover, Random Forest algorithm had the least prediction error in a study related to the utilization of machine learning methods for pregnancy and childbirth risk management in Russia [40]. However, LASSO logistic regression achieved the best performance in predicting the occurrence of pregnancy induced hypertension in first trimester in a study conducted in China [41].

This study also identified the key influencing factors for unintended pregnancy using Information Gain in Senegal. The top 10 predictors that are highly associated with unintended pregnancy in Senegal are total birth, currently residing with husband/partner, respondent’s education level, number of living children, husband/partner’s occupation, residence type, respondent can refuse sex and intention of contraceptive use. A similar study conducted in Ethiopia regarding unintended pregnancy prediction revealed total birth, respondent’s education and refusal sex as important features for unintended pregnancy which aligns with our study [19]. In case of chi square association, region, wealth index, number of living children, total birth, respondent’s occupation and contraceptive use became significantly associated in our study and a similar study in Bangladesh also had the same findings [7]. Respondent’s age, respondent’s education, husband’s education level, age at first cohabitation became insignificant in chi square association test which differs with the above-mentioned study of Bangladesh. Moreover, logistic regression was used to identify the factors influencing the unintended pregnancy in 29 Sub-Saharan African countries where it was revealed that residence, respondent’s education level and contraceptive intention were significant features for unintended pregnancy which agreed with our study [42]. Associated factors of unintended pregnancy among six South Asian countries using multivariable logistic regression where it was found that currently residing with husband, respondent’s education level, number of living children, residence type and intention of contraceptive use were significant features for unintended pregnancy and our study also agrees with it [3].

The research demonstrates that machine learning techniques can effectively pinpoint predictive factors associated with unintended pregnancies. These methods prove valuable in identifying the most critical indicators for predicting unplanned pregnancies in Senegal. Our model holds promise for addressing the significant public health challenge of detecting and addressing unintended pregnancies. These predictions support the provision of comprehensive services and extended working hours for women. By enhancing predictive modeling, we can improve the quality of medical care and boost maternal survival rates. Consequently, the prediction models for unintended pregnancy developed in our study could play a significant role in identifying women at risk of unplanned pregnancies and implementing effective supportive interventions, such as training or information dissemination. This approach may reduce misunderstandings by offering quantitative, objective, and research-based models for risk classification, prediction, and care planning.

### Limitation

This study has several limitations. The predictive model cannot access additional information about other associated parameters when it is constructed using DHS cross-sectional data. The combination of these variables may improve AUC and predicted accuracy. But this research demonstrates that machine learning algorithms can forecast unintended births based on broad risk variables, which can aid in the creation of treatments to enhance planned pregnancies and family planning among Senegalese married couples. Moreover, machine learning model’s result lacks a coefficient and odds ratio, in contrast to the statistical model, making it difficult to assess the degree and direction to which different factors influence the outcome.

## Conclusion

In this study, we evaluated six machine learning algorithms to predict the likelihood of a woman experiencing an unintended pregnancy. Random Forest Classifier produced the best results and the quite accurate classification among the algorithms tested for predicting unplanned pregnancy in Senegalese women. By using Information Gain in order to select the best features related to unintended pregnancy, it was revealed that total birth, currently residing with husband/partner, respondent’s education level, number of living children, husband/partner’s occupation, residence type, respondent can refuse sex and intention of contraceptive use were the significant predictors of unintended pregnancy. In order to improve key policy initiatives, this paper highlights the application of machine learning algorithms to forecast and better understand the most relevant unplanned pregnancy predictor variables.

## Data Availability

The dataset, analyzed in this study, was obtained from the open source of Official Website of DHS program (Weblink: https://www.dhsprogram.com/Data/). The Senegal Demographic and Health Survey 2023 dataset was availed from website for this study (Link: https://dhsprogram.com/methodology/survey/survey-display-611.cfm)

## Acknowledgements

Not applicable

## Authors’ contributions

MRKM, SAT and GK generated the study idea, formed the research team, and designed the study and conducted the initial literature review. SAT wrote the introduction, SAT and GK wrote the methodology and MRKM analyzed the data. MRKM wrote the initial draft of the manuscript. NS supervised the study, reviewed the manuscript for intellectual purposes while MRKM, SAT and GK amended the manuscript according to the feedback and created the final draft of the manuscript. All authors contributed to the intellectual content of the manuscript and read and approved the final version of the manuscript.

## Ethics approval and consent to participate

As the data was obtained from an open source of Official Website of DHS (https://dhsprogram.com/), the ethical approval and consent to participate are not applicable for this study.

## Consent for publication

As the study does not contain any individual and institutional data in any form (including any individual details, images, or videos), the consent for publication is not applicable for this study.

